# Hypoxia as a predictor of mortality among patients admitted with COVID-19 disease in three referral hospitals in Kenya, October 2020 to December 2021

**DOI:** 10.1101/2024.10.17.24315667

**Authors:** Anthony Waruru, Jonesmus Mutua Wambua, Frank Victor Otieno, James Mbai, Mary Mwangome, Peninah Munyua, Wanjiru Waruiru, Sasi Jonnalagadda, Carol Ngunu, Annastacia K. Muange, James Otieno, Dickens Onyango, Nelly Muturi, Anne Njoroge

**Author notes:** **Correspondence**: Anthony Waruru. **Funding**: This publication was made possible by support from the American Rescue Plan Act of 2021 and the U.S. President’s Emergency Plan for AIDS Relief (PEPFAR) through cooperative agreements [GH002338, GH002266, & GH002339] from the U.S. Centers for Disease Control and Prevention (CDC), Division of Global HIV &TB (DGHT).

## Abstract

**Introduction:** Using peripheral oxygen saturation (SpO_2_) measurement is a cost-effective and reliable approach to assess the need for oxygen supplementation in patients admitted with coronavirus disease 2019 (COVID-19). Patients who have a SpO_2_ level of <95% are considered hypoxic and, per COVID-19 management guidelines, should receive oxygen supplementation. We sought to determine whether hypoxia at admission predicted adverse COVID-19 outcomes including mortality among patients admitted with COVID-19 disease in Kenya.

**Methods:** The study was a cross-sectional retrospective medical chart review of patients hospitalized with COVID-19 between October 1, 2020, and December 31, 2021 in three purposively selected health facilities in Kilifi, Nairobi and Kisumu. We explored the differences in proportions of categorical variables using the χ^2^ test and assessed predictors (selected *a priori*) of mortality among patients with hypoxia using Cox proportional hazards models. Using the Kaplan-Meier method, we also computed survival probabilities by hypoxia status for patients on room air or oxygen supplementation and produced survival graphs.

**Results:** Of the 1,124 COVID-19 patients, 94.8% had documented SpO_2_ measurements at admission, and 81.4% were found to have hypoxia, with 39.9% of hypoxic patients not exhibiting dyspnea. Hypoxic patients compared to those with normal oxygen saturation levels were significantly older (60+ years: 44.6 vs. 24.4%) and had a higher prevalence of dyspnea (60.1 vs. 36.9%), higher pulse rate (38.2 vs. 24.6%), and hypertension (40.4 vs. 25.8%), p<0.001. Oxygen supplementation was provided to only 68.6% of hypoxic patients. Mortality was notably higher in hypoxic patients versus those with normal SpO_2_ (38.0% vs. 13.6%, p<0.001), with hypoxia being a key predictor of death. Hypoxic, older patients (≥60 years), and those with dyspnea had a higher risk of death (adjusted hazard ratio: 1.9 [95% confidence interval (CI):1.2–2.8], 1.8 [95% CI 1.3-2.6] and 1.5 [95% CI 1.2-2.0], respectively). Regardless of dyspnea or oxygen supplementation, survival probabilities were worse for hypoxic patients (p<0.001).

**Conclusions:** Hypoxia was prevalent among hospitalized COVID-19 patients, even without respiratory distress symptoms. These findings underscore the importance early identification and management of hypoxia in COVID-19 patients, thereby guiding clinical care and improving outcomes, particularly for older or sicker patients.

**Author summary:** Hypoxia (low oxygen levels) is a common complication in severe COVID-19 and can be a key predictor of poor outcomes, including death. In low-resource settings like Kenya, the availability of advanced diagnostic tools is limited, and simple methods like peripheral oxygen saturation (SpO_2_) measurements are crucial for early identification and management of hypoxia. Despite the importance of oxygen supplementation for hypoxic patients, data on how hypoxia at admission correlates with COVID-19 outcomes in sub-Saharan Africa is limited. We conducted a cross-sectional retrospective study of 1,124 COVID-19 patients hospitalized between October 2020 and December 2021 in three Kenyan counties. We found that 81.4% of patients had hypoxia at admission, but 39.9% of these patients did not exhibit signs of respiratory distress (dyspnea). Hypoxia at admission was associated with higher mortality, especially among older patients (≥60 years) and those with comorbidities, such as hypertension. Survival probabilities for hypoxic patients were significantly lower, regardless of whether they showed signs of dyspnea or received oxygen supplementation. Hypoxia is highly prevalent among hospitalized COVID-19 patients, even in the absence of respiratory distress, highlighting the need for routine oxygen level screening using simple, cost-effective tools like oxymeters. Early detection and management of hypoxia, especially in older or high-risk patients, is essential to improving clinical outcomes in COVID-19. These findings provide evidence for refining COVID-19 management protocols in resource-limited settings, with an emphasis on early intervention to reduce mortality.

## Introduction

Severe acute respiratory syndrome coronavirus type 2 (SARS-CoV-2) virus primarily targets the respiratory system, often leading to pneumonia and acute respiratory distress syndrome (ARDS). Hypoxia in SARS-CoV-2 infection reflects impaired gas exchange in the lungs and results in decreased oxygen levels in the bloodstream.. Studies underscore a strong correlation between hypoxia and the severity of coronavirus disease (COVID-19), with severely affected patients often exhibiting profound hypoxia [1,2]. This hypoxia has been associated with the activation of inflammatory pathways, potentially contributing to the cytokine storm observed in severe COVID-19 cases [3].

Early in the COVID-19 pandemic, it became evident that patients with hypoxia were at higher risk of adverse outcomes [4–7], highlighting the critical role of oxygen therapy in managing severe COVID-19 cases. The saturation of peripheral oxygen (SpO_2_), measured via pulse oximetry, provides crucial information for patient management, aiding in decisions such as hospitalization, especially for elderly patients [8]. SpO_2_ measurements help to categorize patients as having hypoxemia, defined using a cutoff point, with normal oxygen saturation levels in the blood ranging from 95% to 100% and low peripheral oxygen saturation being lower than 95% [9]. In Kenya, hypoxia during COVID-19 was defined as SpO_2_ ≤ 94% on room air [10].

Since pulse oximetry is quick, noninvasive, and painless, the World Health Organization (WHO) recommended its use in monitoring patients with COVID-19 [11], and provided guidance for the clinical management of COVID-19 patients [12]. Despite its simplicity and non-invasiveness, pulse oximetry was underutilized in resource-limited settings during the pandemic, where oxygen availability was scarce [13,14]. A notable observation during the pandemic was “silent hypoxia,” where patients exhibited low SpO_2_ levels without dyspnea, potentially leading to delayed recognition of hypoxemia if SpO_2_ measurements were not routinely performed [6,15,16]. This condition underscores the importance of SpO_2_ monitoring in assessing COVID-19 severity, as abnormal chest X-ray findings do not always correlate with SpO_2_ levels [17].

Silent hypoxia poses a significant challenge in managing COVID-19 patients, as it may indicate early deterioration, and mortality, even in the absence of respiratory symptoms [18–20]. Patients with pre-existing respiratory or cardiovascular conditions, such as chronic obstructive pulmonary disease, or heart disease, are particularly susceptible to hypoxia and face increased risks during COVID-19 infection [21]. Additionally, non-respiratory non-communicable diseases like obesity have been linked to worse COVID-19 outcomes [22,23].

This paper describes the availability of pulse oximetry and the prevalence of hypoxia among COVID-19 patients hospitalized in Kenya during the pandemic. We characterize the impact of hypoxia on COVID-19 outcomes, including mortality rates among hospitalized patients.

## Methods

### Study setting

The first COVID-19 case in Kenya was reported on 13^th^ March 2020. During the period covered by this study Kenya underwent four distinct waves of SARS-CoV-2 infections, driven by different variants *B.1* (2^nd^ wave: October 2020 to January 2021), *Alpha/Beta* (3^rd^ wave: February to May 2021), *Delta* (4^th^ wave: June to November 2021), and *Omicron* (5^th^ wave: December 2021) variants [24]. As of December 31, 2021, Kenya had recorded 297,155 confirmed COVID-19 cases and 5,381 deaths, resulting in a case-fatality rate of 1.8% [25].

### Study sites

The study was conducted in three purposively selected referral facilities in three counties: Mbagathi County Referral Hospital (Nairobi County), Jaramogi Oginga Odinga Teaching and Referral Hospital (Kisumu County), and Kilifi County Referral Hospital (Kilifi County) . The purposive selection of the three facilities was conducted in collaboration with the Division of Integrated Disease Surveillance and Response (DDSR), Ministry of Health, and was based on the high number of COVID-19 cases reported during the selected study period and the prevalence of HIV in the three counties. Nairobi County has a reported HIV prevalence of 3.8%, Kisumu County 17.5%, and Kilifi County 2.3% [26]. As of the end of 2021, the cumulative COVID-19 caseload was 119,538 in Nairobi, 7,368 in Kisumu, and 6,461 in Kilifi County [27].

### Data collection and abstraction procedures

The study involved a retrospective cohort analysis of inpatient medical records from hospitalized COVID-19 patients in Kenya spanning October 1, 2020 to December 31, 2021. A team of trained research assistants comprising clinical and health records officers enumerated all the inpatients with a diagnosis of COVID admitted to the three facilities in that time period. Medical charts for these patients were retrieved from the health records departments for abstraction. Data were abstracted using a formatted data abstraction tool programmed in Open Data Kit (ODK), <https://opendatakit.org>, and submitted to a central server for processing and analyses. All the data were stripped of all personally identifying information.

### Measures

A case **infected with SARS-CoV-2** was defined as a patient with a positive rapid diagnostic test or PCR test result or who was presumptively managed for COVID-19 based on symptomatic presentation. Clinical signs and symptoms were also abstracted, including fever, cough, and chest tightness,. Chest X-ray findings (normal/abnormal), if available, were also abstracted. Co-morbidities such as diabetes and hypertension were abstracted as present if the patient had reported them, even if blood glucose levels or current blood pressure readings were normal. We defined **saturation of peripheral oxygen** (SpO_2_) as a measure of how much hemoglobin is bound to oxygen compared to how much hemoglobin remains unbound. **Hypoxia** was defined as SpO_2_ less than or equal to 94% on room air at admission [10]. **Fever** was defined as elevated body temperature >37.2°C, and **high respiratory rate** (RR) as ≥30 breaths/minute. The outcome variable categories were in-hospital death, discharge, or referral to another facility. Provision of **supplemental oxygen** was captured after admission, and **dyspnea** was symptomatically assessed and documented as a presentation of the patients with respiratory distress or shortness of breath.

### Analyses

We tested for the differences in distribution using the Pearson Chi-square (χ^2^) test. We assessed predictors for mortality for patients with hypoxia using Cox proportional hazards regression after calculating the time from admission to death in days and censoring the outcome. We included all potential predictors in the crude analyses after testing for the assumption of proportionality using the Schoenfield test, but only variables with at least 90% completeness and a significance level of p<0.1 were included in the multivariable model. In the multivariable model, we controlled for oxygen saturation, age (in years), hypertension, diabetes, cardiovascular disease, and dyspnea. We additionally calculated survival probabilities by hypoxia status at admission for patients on room air and oxygen supplementation using the Kaplan–Meier method, and produced survival graphs by hypoxia status.

### Ethical approvals

This activity was reviewed by CDC, deemed not research, and was conducted consistent with applicable federal law and CDC policy.* It was also reviewed by the University of California San Fransico’s Institutional Review Board (UCSF-22-37353) and AMREF’s Ethical and Scientific Review Committee (AMREF-ESRC P1233/2022), the University of California, San Francisco Human Research Protection Program Institutional Review Board (IRB approval number 22-37353), and ethical review committees from the three participating hospitals. The Kenya National Commission for Science, Technology, and Innovation (NACOSTI) permitted this study (License No: NACOSTI/P/22/19903). Due to the study’s retrospective nature and the use of de-identified data, the requirement for individual patient consent was waived.

*45 C.F.R. part 46.102(l)(2), 21 C.F.R. part 56; 42 U.S.C. Sect. 241(d); 5 U.S.C. Sect. 552a; 44 U.S.C. Sect. 3501 et seq.

## Results

### Patient characteristics, clinical presentation, and SpO_2_

Among 1,124 patients hospitalized with COVID-19, 1,066 (94.8%) had documented SpO_2_ measurements at admission, of whom 868 (81.4%) had hypoxia. Hypoxic patients compared to those with normal oxygen saturation levels were significantly older (60+ years: 44.6 vs. 24.4%) and had a higher prevalence of dyspnea (60.1 vs. 36.9%), higher pulse rate (38.2 vs. 24.6%), and hypertension (40.4 vs. 25.8%) (p<0.001). Among patients with hypoxemia, about 2 in every 5, 346 (39.9%) did not have dyspnea p<0.001. A higher proportion of patients with hypoxemia died compared to those with normal SpO_2_ (38.0% vs 13.6%) p<0.001 (Table 1).

**Table 1:** Patients’ demographic and clinical characteristics by hypoxia and mortality, Kenya 2020-21.

| Characteristics* | Total | Hypoxia<sup>†</sup>, n(%) | Normal SpO<sub>2</sub> n(%) | p-value<sup>§</sup> |
| --- | --- | --- | --- | --- |
| **Total** | **1066** | **868 (81.4)<sup>‡</sup>** | **198 (18.6)<sup>‡</sup>** |  |
| **Sex (n=1062)** |  |  |  | 0.065 |
| Male | 567 (53.4) | 474 (54.7) | 93 (47.4) |  |
| Female | 495 (46.6) | 392 (45.3) | 103 (52.6) |  |
| **Age in years (n=1063)** |  |  |  | <0.001 |
| <40 | 245 (23.0) | 152 (17.6) | 93 (47.2) |  |
| 40 – 59 | 384 (36.1) | 328 (37.9) | 56 (28.4) |  |
| ≥60 | 434 (40.8) | 386 (44.6) | 48 (24.4) |  |
| **Occupation (n=665)** |  |  |  | 0.934 |
| Formal | 273 (41.1) | 221 (41.1) | 52 (40.9) |  |
| Informal | 379 (57.0) | 307 (57.1) | 72 (56.7) |  |
| Other | 13 (2.0) | 10 (1.9) | 3 (2.4) |  |
| **County (n=1066)** |  |  |  | 0.297 |
| Kilifi | 146 (13.7) | 119 (13.7) | 27 (13.6) |  |
| Kisumu | 533 (50.0) | 443 (51.0) | 90 (45.5) |  |
| Nairobi | 387 (36.3) | 306 (35.3) | 81 (40.9) |  |
| **HIV status (n=99)** |  |  |  | 0.936 |
| Positive | 88 (88.9) | 65 (89.0) | 23 (88.5) |  |
| Negative | 11 (11.1) | 8 (11.0) | 3 (11.5) |  |
| **Temperature (n=583)** |  |  |  | 0.990 |
| Elevated body temperature (>37.2 °C) | 45 (7.7) | 36 (7.7) | 9 (7.7) |  |
| Normal (36.5-37.2 °C) | 538 (92.3) | 430 (92.3) | 108 (92.3) |  |
| **Cough (n=1066)** |  |  |  | <0.001 |
| Yes | 269 (25.2) | 190 (21.9) | 79 (39.9) |  |
| No | 797 (74.8) | 678 (78.1) | 119 (60.1) |  |
| **Chest tightness (n=1066)** |  |  |  | 0.177 |
| Yes | 584 (54.8) | 467 (53.8) | 117 (59.1) |  |
| No | 482 (45.2) | 401 (46.2) | 81 (40.9) |  |
| **Hypertension (n=1066)** |  |  |  | <0.001 |
| Yes | 402 (37.7) | 351 (40.4) | 51 (25.8) |  |
| No | 664 (62.3) | 517 (59.6) | 147 (74.2) |  |
| **Diabetes (n=1066)** |  |  |  | 0.110 |
| Yes | 251 (23.5) | 213 (24.5) | 38 (19.2) |  |
| No | 815 (76.5) | 655 (75.5) | 160 (80.8) |  |
| **Pulse rate (PR) (n=970)** |  |  |  | <0.001 |
| Low PR (<60 bpm) | 35(3.6) | 34 (4.3) | 1 (0.5) |  |
| Normal PR (60-100 bpm) | 590(60.8) | 450 (57.5) | 140 (74.9) |  |
| High PR (>100 bpm) | 345(35.6) | 299 (38.2) | 46 (24.6) |  |
| **Cardiovascular issues** |  |  |  | 0.281 |
| With issues | 1045 (98.0) | 849 (97.8) | 196 (99.0) |  |
| No issues | 21 (2.0) | 19 (2.2) | 2 (1.0) |  |
| **Dyspnea (n=1066)** |  |  |  | <0.001 |
| Dyspnea | 595 (55.8) | 522 (60.1) | 73 (36.9) |  |
| None | 471 (44.2) | 346 (39.9) | 125 (63.1) |  |
| **Oxygen supplementation (n=1066)** |  |  |  | <0.001 |
| Yes | 644(60.4) | 595(68.6) | 49(24.8) |  |
| No | 422(39.6) | 273(31.5) | 149(75.3) |  |
| **Patient Outcome (n=1066)** |  |  |  | <0.001 |
| Dead | 357(33.5) | 330 (38.0) | 27 (13.6) |  |
| Discharged | 680(63.8) | 516 (59.4) | 164 (82.8) |  |
| Referred out | 29(2.7) | 22 (2.5) | 7 (3.5) |  |

\*Some variables had missing values. The number of included records is indicated against the variable names.
\*Hypoxia was defined as SpO<sub>2</sub> reading of $\leq 94\%$ on room air measured during admission.
‡Row percentages
§Chi-square p-values, significance at $p < 0.05$

### Dyspnea and hypertension in relation to patient outcomes by SpO_2_ at admission

Among all COVID-19 patients without dyspnea, hypoxic patients had three-fold more deaths than those with normal SpO_2_ saturation (29.8% vs. 9.6%), p<0.001. Among all COVID-19 patients with dyspnea, hypoxic patients had two-fold more deaths than those with normal SpO_2_ saturation (43.5% vs. 20.6%), p=0.002 (Figure 1).

**Figure 1:**
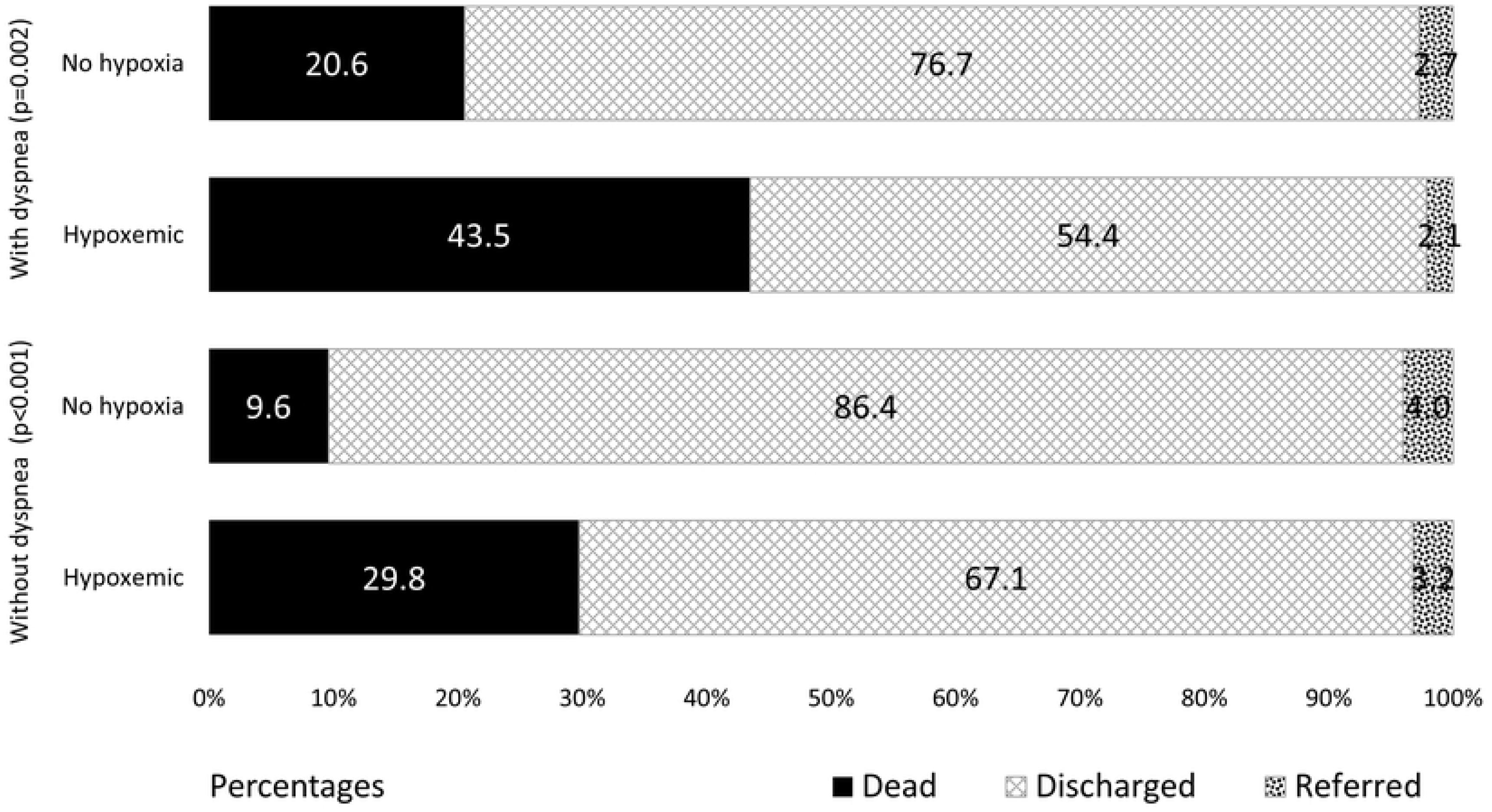
Hypoxia and outcomes for COVID-19 patients with and without dyspnea, Kenya 2020-21. Describes the outcomes for COVID-19 patients by dyspnea.

Among all COVID-19 patients, those with hypoxia had a higher proportion of deaths than those with normal SpO_2_ saturation (38.0% vs. 13.6%), p<0.001. There were more deaths among COVID-19 patients with hypertension and hypoxia compared to hypertensive patients with normal SpO_2_ saturation (42.5% vs. 7.8%), p<0.001. For COVID-19 patients without hypertension, mortality was higher among patients who had hypoxia (35.0%) compared to patients with normal SpO_2_ saturation (15.6%), p<0.001 (Figure 2).

**Figure 2:**
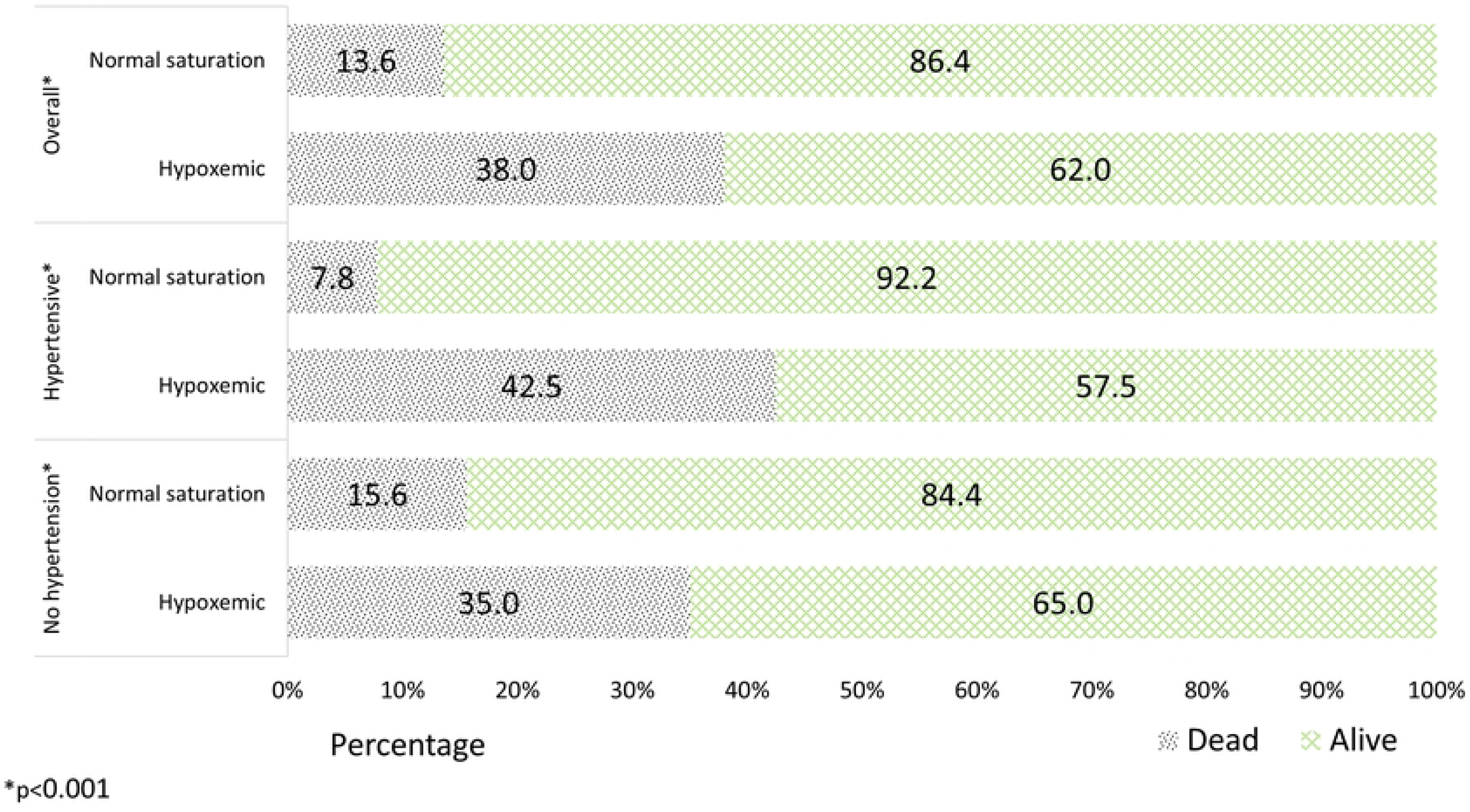
Hypoxia and mortality for all patients with and without hypertension, Kenya 2020-21. Describes mortality proortions for all patients with and without hypertension.

### Influence of SpO_2_ at admission on survival probability

Whether on oxygen supplementation or room air, survival probabilities were better for patients who did not have hypoxia on admission. Survival for non-hypoxic COVID-19 patients on room air plateaued from about two weeks of hospitalization, while survival for hypoxic patients continued to worsen at a more rapid pace (Figure 3a). Both hypoxic and non-hypoxic patients had similar declining survival rates beyond a month of hospitalization. Half of the hypoxic patients on room air survived up to the 20^th^ day, and over half of the non-hypoxic patients on room air survived over the 50^th^ day. For patients on supplemental oxygen (Figure 3b), survival probabilities rapidly worsened, and by around 10 days, over half of the hypoxic patients on admission were dead, compared to non-hypoxic patients on admission who survived a little longer.

**Figure 3:**
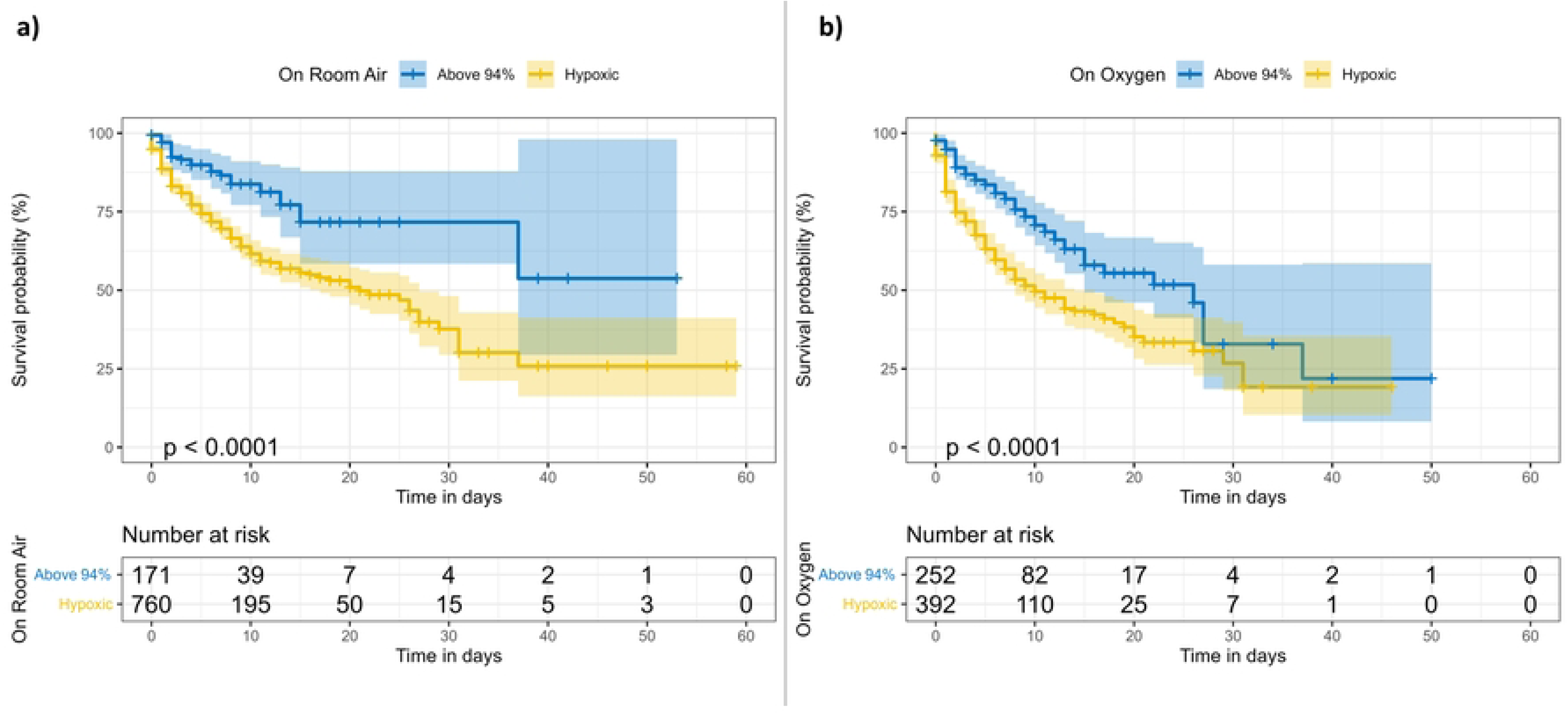
Survival probabilities for COVID-19 patients on room air (a) and on oxygen supplementation (b), by oxygen saturation levels, Kenya 2020-21. Shows Kaplan meier survival probabilities for patients who were not on oxygen supplementation (room air) and those who were on oxygen supplementation.

### Predictors of mortality among patients with hypoxia

Out of 1,123 patients admitted with COVID-19, 1122 recorded time-to-outcome data, and 868 experienced hypoxia. The median duration of admission was 6 days, interquartile range (IQR) 2-10. Among the 1066 patients with hypoxia data, 357 (33.5%) died. Mortality was significantly higher in hypoxic patients (38.0%) compared to those with normal oxygen levels (13.6%) (p<0.001). Lower mortality rates were observed in non-hypoxic patients, females, those under 40, those with formal employment, HIV negative individuals, and those without hypertension, diabetes, cardiovascular issues, or dyspnea. Crude proportional hazards analysis identified hypoxia, age, hypertension, diabetes, cardiovascular issues, and dyspnea as significant factors affecting survival (p<0.05). In the adjusted model, hypoxia (adjusted hazard ratio (aHR) = 1.9 [95% confidence interval (CI):1.2–2.8]), older age (60 + years) (aHR) = 1.8 [95% CI:1.3–2.6]), and dyspnea (aHR 1.5 [95% CI: 1.2–2.0]) were associated with a higher risk of death (Table 2).

**Table 2:**
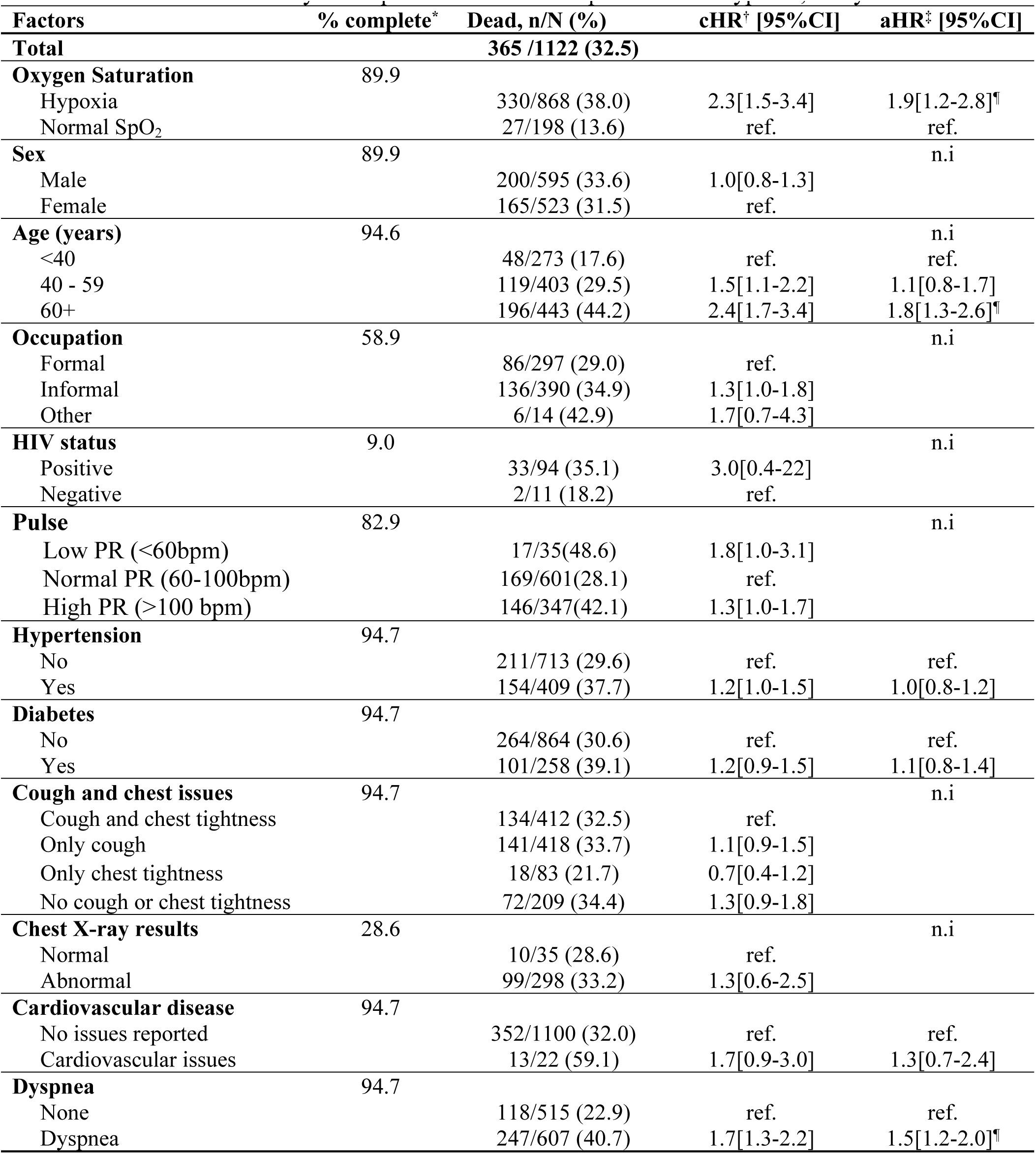
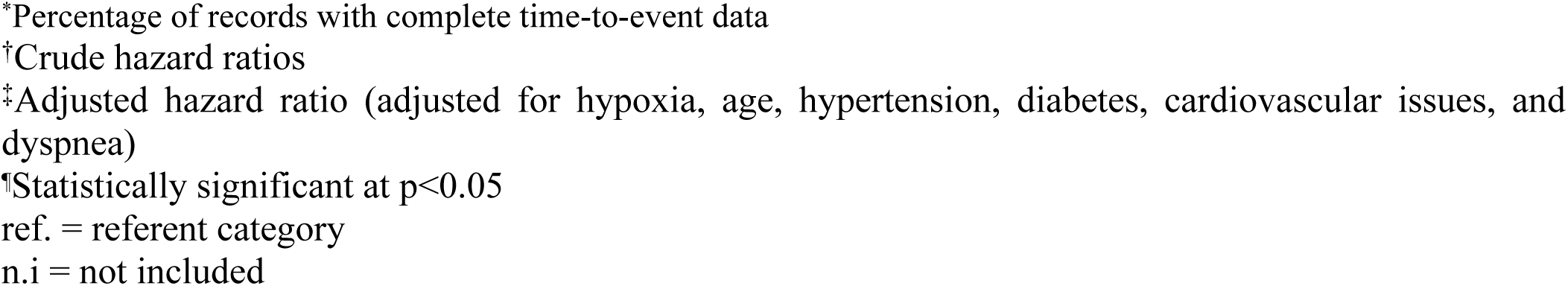
Predictors of mortality in hospitalized COVID-19 patients with hypoxia, Kenya 2020-21.

| Factors | % complete* | Dead, n/N (%) | cHR<sup>†</sup> [95%CI] | aHR<sup>‡</sup> [95%CI] |
| --- | --- | --- | --- | --- |
| **Total** |  | **365 /1122 (32.5)** |  |  |
| **Oxygen Saturation** | 89.9 |  |  |  |
| Hypoxia |  | 330/868 (38.0) | 2.3[1.5-3.4] | 1.9[1.2-2.8]<sup>¶</sup> |
| Normal SpO<sub>2</sub> |  | 27/198 (13.6) | ref. | ref. |
| **Sex** | 89.9 |  |  | n.i |
| Male |  | 200/595 (33.6) | 1.0[0.8-1.3] |  |
| Female |  | 165/523 (31.5) | ref. |  |
| **Age (years)** | 94.6 |  |  | n.i |
| <40 |  | 48/273 (17.6) | ref. | ref. |
| 40 - 59 |  | 119/403 (29.5) | 1.5[1.1-2.2] | 1.1[0.8-1.7] |
| 60+ |  | 196/443 (44.2) | 2.4[1.7-3.4] | 1.8[1.3-2.6]<sup>¶</sup> |
| **Occupation** | 58.9 |  |  | n.i |
| Formal |  | 86/297 (29.0) | ref. |  |
| Informal |  | 136/390 (34.9) | 1.3[1.0-1.8] |  |
| Other |  | 6/14 (42.9) | 1.7[0.7-4.3] |  |
| **HIV status** | 9.0 |  |  | n.i |
| Positive |  | 33/94 (35.1) | 3.0[0.4-22] |  |
| Negative |  | 2/11 (18.2) | ref. |  |
| **Pulse** | 82.9 |  |  | n.i |
| Low PR (<60bpm) |  | 17/35(48.6) | 1.8[1.0-3.1] |  |
| Normal PR (60-100bpm) |  | 169/601(28.1) | ref. |  |
| High PR (>100 bpm) |  | 146/347(42.1) | 1.3[1.0-1.7] |  |
| **Hypertension** | 94.7 |  |  |  |
| No |  | 211/713 (29.6) | ref. | ref. |
| Yes |  | 154/409 (37.7) | 1.2[1.0-1.5] | 1.0[0.8-1.2] |
| **Diabetes** | 94.7 |  |  |  |
| No |  | 264/864 (30.6) | ref. | ref. |
| Yes |  | 101/258 (39.1) | 1.2[0.9-1.5] | 1.1[0.8-1.4] |
| **Cough and chest issues** | 94.7 |  |  | n.i |
| Cough and chest tightness |  | 134/412 (32.5) | ref. |  |
| Only cough |  | 141/418 (33.7) | 1.1[0.9-1.5] |  |
| Only chest tightness |  | 18/83 (21.7) | 0.7[0.4-1.2] |  |
| No cough or chest tightness |  | 72/209 (34.4) | 1.3[0.9-1.8] |  |
| **Chest X-ray results** | 28.6 |  |  | n.i |
| Normal |  | 10/35 (28.6) | ref. |  |
| Abnormal |  | 99/298 (33.2) | 1.3[0.6-2.5] |  |
| **Cardiovascular disease** | 94.7 |  |  |  |
| No issues reported |  | 352/1100 (32.0) | ref. | ref. |
| Cardiovascular issues |  | 13/22 (59.1) | 1.7[0.9-3.0] | 1.3[0.7-2.4] |
| **Dyspnea** | 94.7 |  |  |  |
| None |  | 118/515 (22.9) | ref. | ref. |
| Dyspnea |  | 247/607 (40.7) | 1.7[1.3-2.2] | 1.5[1.2-2.0]<sup>¶</sup> |

\*Percentage of records with complete time-to-event data †Crude hazard ratios ‡Adjusted hazard ratio (adjusted for hypoxia, age, hypertension, diabetes, cardiovascular issues, and dyspnea) §Statistically significant at $p < 0.05$ ref. = referent category n.i = not included

## Discussion

This study highlights the critical role of hypoxia in determining the outcomes of patients hospitalized with COVID-19. Notably, 8 out of 10 patients hospitalized with COVID-19 had hypoxia at admission, a condition that was associated with significantly higher mortality rates compared to those with normal oxygen saturation levels. The prevalence of hypoxia was particularly higher among older patients and those with clinical symptoms consistent with COVID-19, such as cough, chest tightness, low pulse rate, dyspnea, and underlying comorbidities like hypertension and HIV infection. This relationship suggests that symptomatic COVID-19 disease may have a consistent prognosis even in the absence of diagnostic testing, a finding supported by similar studies [5]. Interestingly, about 40% of our patients experienced silent hypoxia, a proportion that aligns with estimates from other studies, which range between 4.8% and 65% [5]. This emphasizes the importance of pulse oximetry at triage or admission, even for patients without apparent respiratory symptoms, to enable timely supplementation and other interventions that can improve outcomes.

Mortality in our study was over twice as high among patients with hypoxia compared to those with normal SpO_2_ saturation (38.0% vs. 13.6%). This underscores the severe consequences of oxygen deprivation in COVID-19, corroborating existing literature that identifies hypoxia as a key predictor of poor prognosis in respiratory infections [18,19,28]. Moreover, older hypoxic patients had twice the risk of death compared with patients under 40, which is consistent with global findings [29]. While hypertension was not independently associated with mortality in our study, it is generally recognized as a risk factor for severe COVID-19 and poor outcomes [30]. SpO_2_ has emerged as a crucial predictor of survival in COVID-19 patients [20]. Our findings suggest that normal SpO_2_ levels are associated with better survival probabilities, while hypoxia, along with age and dyspnea, independently predicts a higher risk of death. Therefore, oxygen supplementation is essential for hypoxic patients, as studies have shown that higher SpO_2_ levels after supplementation are linked to reduced mortality [5], and improved survival probabilities [19]. The observation that patients with hypoxia had over three times the mortality rate underscores the importance of assessing oxygen saturation in all COVID-19 patients, regardless of their symptomatic presentation.

Oxygen supplementation is recommended for hypoxic patients. However, only for only 3 out of 5 (68.6%) of hypoxic patients in our study received oxygen supplementation, indicating insufficient management in our setting. The national guidelines recommend oxygen supplementation for patients with SpO_2_ ≤94%, but a more strict target of 92–96% might have been beneficial [31], given that the Kenyan cutoff was less stringent. We also lacked data on whether follow-up SpO_2_ readings were taken after initial drops by 3-5%, which is a recommended practice, as has been suggested by Galwankar et al. [32]. Community-level SpO_2_ monitoring, such as providing oximeters to households [34], could be beneficial for ongoing patient management post-discharge, though we did not follow patients after discharge.

Our study had some limitations. First, we lacked COVID-19 diagnostic information for some cases, relying instead on clinician documentation based on symptomatic presentation. Additionally, SpO_2_ data were missing for 5.2% of records, but this low proportion is unlikely to bias our findings. The relatively stringent definition of hypoxemia used in this study may have led to the misclassification of some patients as having silent hypoxemia, though we adhered to national guidelines. The study’s findings may not be generalized to the broader Kenyan population, as the sites were purposefully selected. We also did not track patient outcomes of COVID-19 patients post-discharge. Finally, we could not determine whether hypertensive patients had a history of hypertension or elevated blood pressure before admission. Despite these limitations, our study provides valuable insights into the utility of SpO_2_ monitoring for the clinical management of hospitalized COVID-19 patients.

Our study suggests that while comorbid conditions like hypertension, diabetes, and cardiovascular disease are important, they did not significantly alter mortality risk after adjusting for hypoxia, age, and dyspnea. This indicates that hypoxia and age are the predominant drivers of mortality in hospitalized COVID-19 patients, with other comorbidities playing a secondary role. The frequent occurrence of hypoxia among hospitalized COVID-19 patients, often without accompanying dyspnea, underscores the importance of using SpO_2_ measurements to guide hospitalization decisions and improve outcomes, particularly for older or more critically ill patients. Finally, the higher mortality among patients with silent hypoxia highlights the need for regular SpO_2_ monitoring to enhance patient outcomes, especially for those with comorbidities.

## Data Availability

The data underlying the results presented in the study are available from the head Division of Disease Surveillance and Response, email: [contact authors]

## Acknowlegements

We thank the Kenyan Ministry of Health, the Division of Disease Surveillance and Response (DDSR), the National AIDs and STIs Control Programme, VL laboratories, implementing partners, and patients. This work was supported by the President’s Emergency Plan for AIDS Relief (PEPFAR) through the Centers for Disease Control and Prevention (CDC).

## Authors contributions

AW analyzed the data and drafted the initial and subsequent drafts of the manuscript, JMW contributed to the analyses. AN, DO, MM, and WW contributed substantially to protocol development, data collection, and critical review of the manuscript. All authors read and approved the final version of the manuscript.

## Disclaimer

The findings and conclusions in this manuscript are those of the authors and do not necessarily represent the official position of the funding agencies.

